# The Role of Behavioral and Cognitive Disorders in Determining Post-Discharge Residential Status from Mental Health Facilities: Evidence from SAMHSA MH-CLD Data

**DOI:** 10.1101/2025.08.01.25332815

**Authors:** Eden Moges, Norma Rochez, William Peycha, Aditya Chakraborty

## Abstract

**Background:** Behavioral and cognitive disorders are significant public health concerns, often influencing housing stability and residential outcomes. Access to stable housing is a key determinant of mental health recovery, yet disparities persist across demographic and socioeconomic groups. This study examines the association between behavioral and cognitive disorders and residential status at discharge from mental health facilities.

**Methods:** Using the 2022 Mental Health Client-Level Data from the Substance Abuse and Mental Health Services Administration (SAMHSA), this cross-sectional study analyzed the relationship between primary behavioral and cognitive disorders, such as ADHD, Oppositional Defiant Disorder (ODD), and Personality Disorders, and residential status upon discharge. Descriptive statistics were used to summarize demographic characteristics, while logistic regression models were employed to assess associations between disorders and residential outcomes, adjusting for age, race, employment status, education, substance use, and gender.

**Results:** Of the 6,957,919 individuals analyzed, 3.7% experienced homelessness, 11.4% resided in alternative settings (e.g., foster care, residential treatment, correctional facilities), and 84.9% lived in private residences. Individuals with behavioral and cognitive disorders were significantly less likely to reside in private residences compared to those without such disorders (AOR = 0.876, 95% CI: 0.866– 0.887, p < 0.0001). Additional factors such as substance use (AOR = 0.798, 95% CI: 0.787–0.809, p < 0.0001), unemployment (AOR = 2.60, 95% CI: 2.578–2.624, p < 0.0001), and lower education levels (AOR = 1.543, 95% CI: 1.526–1.561, p < 0.0001) further predicted non-private residential outcomes. Racial and gender disparities were also looked at, with African Americans being 34% more likely to experience non-private residential placements compared to white individuals (AOR = 1.340, 95% CI: 1.323–1.358, p < 0.0001). Additionally, males had greater odds of experiencing homelessness or alternative housing compared to females (AOR = 1.192, 95% CI: 1.176–1.208, p < 0.0001).

**Conclusion:** The findings highlight the intersection of mental health disorders, socioeconomic factors, and housing stability. Individuals with behavioral and cognitive disorders face significant barriers to stable housing, exacerbated by substance use, unemployment, and racial disparities. These results reinforce the need for integrated mental health and housing interventions, with a focus on culturally competent policies to improve residential stability for vulnerable populations.

## INTRODUCTION

Mental health is a complex and multifactorial concept influenced by a wide range of biological, psychological, and social factors. Among these, behavioral and cognitive disorders, such as Attention-Deficit/Hyperactivity Disorder (ADHD), Oppositional Defiant Disorder (ODD), Conduct Disorder, Personality Disorders, and Pervasive Developmental Disorder (PDD), represent a group of conditions that significantly impair daily functioning and long-term life outcomes. These disorders often manifest in childhood and adolescence, but their effects can persist into adulthood, leading to enduring challenges in employment, social integration, and independent living. One of the most significant issues faced by individuals with these disorders is securing and maintaining stable housing, a problem that remains a persistent concern in both clinical and public health contexts (Pawliczuk et al., 2018).

Research consistently demonstrates that individuals with behavioral and cognitive disorders are at a heightened risk of housing instability. These individuals often experience difficulties in forming and maintaining stable relationships, managing financial obligations, and navigating complex systems like employment and social services (Wu et al., 2021). As a result, they are more likely to experience homelessness or be placed in institutional care settings such as foster homes, residential treatment facilities, or correctional institutions. The transition from institutional settings to independent living is particularly challenging, as individuals with these disorders often lack the social skills, coping mechanisms, and resources necessary to maintain stable housing. Studies have shown that children and youth with behavioral disorders are overrepresented in residential institutions, and their experiences in these environments often leave them vulnerable to housing instability later in life (Kwan & Rickwood, 2015).

This study aims to examine the relationship between residential status at discharge and the prevalence of behavioral and cognitive disorders using the Mental Health Client-Level Data (MH-CLD) 2022, provided by the Substance Abuse and Mental Health Services Administration (SAMHSA, 2021). The MH-CLD dataset includes detailed information on mental health diagnoses, services provided, and demographic characteristics, making it a valuable resource for understanding how different diagnoses, particularly behavioral and cognitive disorders, are associated with residential outcomes upon discharge. This study seeks to explore how these disorders contribute to disparities in housing stability, particularly in relation to age, race, and socio-economic factors.

Prior research has highlighted the critical role of behavioral disorders in shaping housing outcomes. Individuals with these conditions are disproportionately likely to experience homelessness or be placed in non-private living arrangements. The intersection of behavioral disorders with other socio-economic factors, such as education, employment status, and substance use, complicates the likelihood of achieving stable housing. For instance, substance use is a common comorbidity among individuals with behavioral disorders, and it is strongly correlated with housing instability. The absence of stable housing exacerbates mental health symptoms, creating a vicious cycle where individuals with behavioral disorders face increasing difficulties in securing permanent, stable housing. (World Health Organization, 2022).

The MH-CLD dataset is critical for evaluating National Outcome Measures (NOMs), which include indicators of housing stability, employment, access to mental health services, and clinical improvements. By examining these outcomes, this study aims to shed light on how behavioral and cognitive disorders affect residential status, revealing the broader implications for public health and housing policy. Previous studies have also demonstrated that individuals with mental health disorders are often faced with substantial barriers to housing, and this problem is particularly acute among vulnerable populations, including racial minorities and individuals with lower socio-economic status.

Culturally competent mental health care is essential in addressing the disparities in housing and service utilization (Bhui et al., 2007). Ethnic minorities, for example, often experience additional barriers such as cultural stigma surrounding mental health and limited access to mental health services, which further complicates their ability to maintain stable housing (Saasa et al., 2021). Addressing these disparities requires integrating mental health services with housing and social support systems, recognizing the importance of social determinants of health, including housing. (World Health Organization, 2022). The need for culturally adapted mental health services is increasingly recognized, as such approaches have been shown to improve both service utilization and outcomes (Rice & Harris, 2020).

At a global level, the World Health Organization (WHO) has underscored the importance of systemic changes in mental health services, particularly for vulnerable populations. The WHO’s 2022 report called for a holistic approach to care that integrates housing with mental health services, aligning with SAMHSA’s mission to reduce barriers to care and improve accessibility to mental health services. (World Health Organization, 2022). The MH-CLD dataset complements these objectives by providing data-driven insights that inform policies designed to improve both mental health and housing outcomes for individuals with behavioral and cognitive disorders (SAMHSA, 2021).

This research aims to explore patterns in residential outcomes, including homelessness, private residence placements, and alternative living arrangements such as foster care and residential treatment facilities, among individuals with behavioral and cognitive disorders. By understanding these associations, this study seeks to inform policies that integrate mental health care with housing support, ultimately aiming to reduce housing disparities for individuals with mental health conditions. Previous studies have shown that youth with mental health disorders are particularly vulnerable to housing instability, which can hinder their developmental progress and social integration. (Pawliczuk et al., 2018). This study intends to further explore the link between these disorders and housing outcomes, emphasizing the importance of coordinated interventions that address both mental health needs and housing stability.

## METHODS

### Study Design and Data Source

This study employed a cross-sectional design using the Mental Health Client-Level Data (MH-CLD) 2022, provided by the Substance Abuse and Mental Health Services Administration (SAMHSA). The MH-CLD dataset is a nationally representative database that includes comprehensive data on individuals receiving mental health services across State Mental Health Agencies (SMHAs) in the United States. It contains detailed information on participants’ demographics, clinical diagnoses, service utilization, and residential outcomes. Data from this dataset are weighted to account for the complex survey design, nonresponse, and noncoverage, ensuring the generalizability of the results to the broader population of individuals receiving mental health services.

#### Study Population

The initial MH-CLD 2022 dataset included data on 6,957,919 participants. After applying inclusion criteria, the final sample consisted of 3,222,709 individuals. The study focused on participants with complete data on behavioral and cognitive disorders, residential status, and other relevant covariates. Participants were categorized based on behavioral and cognitive disorder status (present vs. absent) and residential status (private vs. non-private residence).

### Measures

#### Behavioral and Cognitive Disorders

Behavioral and cognitive disorders were identified based on participants’ mental health diagnoses, including Attention-Deficit/Hyperactivity Disorder (ADHD), Oppositional Defiant Disorder (ODD), Conduct Disorder, Personality Disorder, and Pervasive Developmental Disorder (PDD).

Individuals diagnosed with one or more of these conditions were categorized as having a behavioral and cognitive disorder.

#### Residential Status

The primary outcome variable, residential status, was classified into a binary variable:

- Private Residence: Individuals discharged to a private residence, including family homes.
- Non-Private Residence: Individuals discharged to homelessness or other institutional settings such as foster care, residential treatment facilities, or correctional institutions.

#### Covariates

Several covariates were included to adjust for potential confounding factors:

1. Gender (GENDER): Coded as male or female.
2. Education Level (EDUC_group): Coded into categories: no diploma/under 12th grade, high school diploma/GED, and beyond highschool
3. Substance Use (SAP_num): Proxy for substance use, based on the number of previous mental health services received.
4. Employment Status (employ_status): Coded as employed or unemployed/not reported.
5. Race/Ethnicity (raceclass): Categorical variable including Hispanic, Native American, Asian, Black, Pacific Islander, White, and Other/Two or more races.
6. Age Group (Age_Group): Coded as 0-17 years, 18–44 years, and 45+
7. Service Site (service_site): Coded as community-based programs or residential treatment facilities.
8. SMISED Status (SMISED): Coded as SMI, SED or at risk for SED, No SMI/SED.
9. Number of Mental Health Diagnoses reported (NUMMHS): Coded by the number of already reported diagnoses, from 0-3.

### Statistical Analysis

Descriptive statistics were used to summarize the demographic characteristics of the study sample and the prevalence of behavioral and cognitive disorders and residential status. To examine associations between behavioral and cognitive disorders, residential status, and covariates, chi-square tests were conducted for categorical variables, while t-tests were applied for continuous variables. Three logistic regression models were performed. The first model, an unadjusted logistic regression, assessed the crude relationship between behavioral and cognitive disorders and residential status. The second model, a multivariable logistic regression, adjusted for covariates, estimated the adjusted odds ratio (AOR) for the likelihood of being discharged to a non-private residence compared to a private residence.

The third model tested interaction terms to explore whether the relationship between behavioral and cognitive disorders and residential status differed by gender, age, education, employment status, race/ethnicity, service site, marital status, or number of mental health services. The odds ratios (ORs) and adjusted odds ratios (AORs) with 95% confidence intervals (CIs) were reported to quantify the strength and direction of associations. Statistical significance was set at p < 0.001 for all tests. All analyses were conducted using SAS software (Version 9.4).

### Ethical Considerations

This observational study used de-identified, publicly available data from the Substance Abuse and Mental Health Services Administration (https://www.samhsa.gov/data/report/2022-mental-health-client-level-data-annual-report), which contain no personal identifiers or protected health information. As such, the study did not involve human subjects as defined by 45 CFR 46, and institutional review board (IRB) approval was not required. The study followed the ethical standards outlined in the Declaration of Helsinki and adheres to the STROBE guidelines for reporting observational studies.

## RESULTS

The final study sample consisted of 3,222,709 participants from the MH-CLD 2022 dataset. Of these, 1,534,121 (47.6%) had been diagnosed with a behavioral and cognitive disorder. The majority of participants were female (61.4%), and the sample was predominantly non-Hispanic White (44.8%), followed by non-Hispanic Black (26.7%), Hispanic (19.5%), and other racial/ethnic groups. The demographic breakdown shows a diverse population, enabling an analysis of how different racial and ethnic groups are affected by behavioral and cognitive disorders in terms of residential stability.

Regarding residential status, 2,045,774 participants (63.5%) were discharged to a private residence, while 1,176,935 participants (36.5%) were discharged to non-private residence, including homelessness or other institutional care settings.

Among those with behavioral and cognitive disorders, 48.3% were discharged to non-private residence, compared to 33.2% of individuals without these disorders, showing a clear disparity in housing stability between these two groups. The average age of participants in the study was 37.4 years, with participants in the non-private residence group being significantly older (mean age of 40.2 years) compared to those in the private residence group (mean age of 35.1 years, p < 0.001). Additionally, participants in the non-private residence group were more likely to report lower education levels, unemployment, and higher substance use compared to those in the private residence group, indicating that these socio-economic factors are closely associated with housing instability in this population.

**Table 1:** Demographic analysis of characteristics of participants by residential status at discharge.

| Variable | Residential Status |  | P-value |
| --- | --- | --- | --- |
|  | Private Residence<br>N (%) | Non-Private Residence<br>N (%) |  |
| Age ( in groups) |  |  |  |
| 0-17 | 719897 (25.25) | 31923 (8.60) | <0.0001 |
| 18-44 | 1278632 (44.84) | 169351 (45.61) |  |
| 45+ | 852897 (29.91) | 169981 (45.78) |  |
| Gender |  |  |  |
| Male | 1282431 (44.49) | 222077 (59.82) | <0.0001 |
| Female | 1569011 (55.03) | 149190 (40.18) |  |
| Race/ethnicity |  |  |  |
| Hispanic | 374331 (13.13) | 36705 (9.89) | <0.0001 |
| Non-Hispanic Native American | 35066 (1.23) | 6247 (1.68) |  |
| Non-Hispanic Asian | 29734 (1.04) | 2663 (0.72) |  |
| Non-Hispanic Black | 539608 (18.92) | 88062 (23.72) |  |
| Non-Hispanic Pacific Island | 4023 (0.14) | 678 (0.18) |  |
| Non-Hispanic White | 1525783 (53.51) | 198488 (53.46) |  |
| Other race/ Two or more | 142207 (4.99) | 15504 (4.18) |  |
| Employment Status |  |  |  |
| Employed | 540894 (18.97) | 32033 (8.63) | <0.0001 |
| Unemployed/not reported | 2310548 (81.03) | 339234 (91.37) |  |
| Education |  |  |  |
| No Diploma/Under 12th | 1273235 (44.65) | 138441(37.29) | <0.0001 |
| Highschool Diploma/GED | 1029251 (36.10) | 170063(45.81) |  |
| Beyond Highschool | 548956 (19.25) | 62763(16.91) |  |
| Substance Abuse Problem |  |  |  |
| Yes | 1195778 (41.94) | 190026 (51.18) | <0.0001 |
| No/ Not Reported | 1655664 (58.06) | 181241 (48.82) |  |
| Service Site |  |  |  |
| Community-Based Program | 2691424 (94.39) | 301040 (81.08) | <.0001 |
| Justice System Institution | 2050 (0.07) | 15710 (4.23) |  |
| Residential Treatment Center | 107980 (3.79) | 26260 (7.07) |  |
| State Psych Hospital | 17720 (0.62) | 10057 (2.71) |  |
| Other Psych inpatient | 32268 (1.13) | 18200 (4.90) |  |
| SMI/SED |  |  |  |
| SMI | 1471774 (51.62) | 242941(65.44) | <0.0001 |
| SED and/or at risk for SED | 501385 (17.58) | 24648 (6.64) |  |
| Not SMI/SED | 742364 (26.03) | 83014 (22.36) |  |
| NUMMHS |  |  |  |
| 0 | 277620 (9.74) | 48049 (12.94) | <0.0001 |
| 1 | 1379342 (48.80) | 182736 (49.22) |  |
| 2 | 841239 (29.50) | 101360 (27.30) |  |
| 3 | 353241 (12.39) | 39122 (10.54) |  |
| Behavioral/Cognitive Disorder |  |  |  |
| Yes | 600672 (21.07) | 59939(16.14) | <0.0001 |
| No | 2250770 (78.93) | 311328 (9.66) |  |

In the unadjusted logistic regression, individuals with a behavioral and cognitive disorder were significantly more likely to be discharged to non-private residence than those without these disorders (OR = 1.52, 95% CI: 1.50–1.54, p < 0.001). This indicates that individuals with these disorders were 52% more likely to experience housing instability compared to those without a behavioral or cognitive disorder. Additionally, participants in the non-private residence group were significantly older (mean age 40.2 years) compared to those in the private residence group (mean age 35.1 years, p < 0.001). These results underscore the significant impact of mental health disorders on housing outcomes and highlight the need for targeted interventions that consider mental health when addressing housing instability.

In the multivariable logistic regression model, after adjusting for key covariates, including age, gender, race/ethnicity, education, employment status, substance use, service site, and SMI/SED status, individuals with a behavioral and cognitive disorder remained significantly more likely to be discharged to non-private residence (AOR = 1.32, 95% CI: 1.30–1.34, p < 0.001). This indicates that, even after accounting for other socio-economic and demographic factors, individuals with these disorders are still at increased risk of housing instability.

Additional covariates, including age and education level, were also found to be significant predictors of residential status. Specifically, older participants and those with higher education levels were less likely to be discharged to non-private residence (AOR for age 45+ = 1.52, 95% CI: 1.504– 1.538, p < 0.001; AOR for beyond high school = 0.772, 95% CI: 0.760–0.783, p < 0.001), reflecting the protective effects of education on housing outcomes. Furthermore, substance use was found to be a significant predictor of residential status, with individuals reporting higher substance use having increased odds of being discharged to non-private residence (AOR = 1.136, 95% CI: 1.120–1.152, p < 0.001).

**Table 2:** Multivariable Logistic regression model demonstrating the association between behavioral/cognitive disorders and residential status outcomes at discharge (OR, 95% CI)

| Variable | AOR (95% CI) | P-value |
| --- | --- | --- |
| Age ( in years) |  |  |
| 0-17 | 0.242 [0.227, 0.258] | <0.0001 |
| 18-44 (ref) | — |  |
| 45+ | 1.521 [1.504, 1.538] |  |
| Gender |  |  |
| Male (ref) | — | <0.0001 |
| Female | 0.573 [0.568, 0.578] |  |
| Race/ethnicity |  |  |
| Hispanic | 0.750 [0.736, 0.764] | <0.0001 |
| Non-Hispanic Native American | 1.330 [1.255, 1.408] |  |
| Non-Hispanic Asian | 0.643 [0.602, 0.686] |  |
| Non-Hispanic Black | 1.088 [1.074, 1.103] |  |
| Non-Hispanic Pacific Island | 1.317 [1.184, 1.464] |  |
| Non-Hispanic White (ref) | — |  |
| Other race/ Two or more | 0.944 [0.925, 0.964] |  |
| Employment Status |  |  |
| Employed (ref) | — | <0.0001 |
| Unemployed | 2.669 [2.641, 2.698] |  |
| Education |  |  |
| No Diploma/Under 12th | 1.073 [1.061, 1.086] | <0.0001 |
| Highschool Diploma/GED | — |  |
| Beyond Highschool | 0.772 [0.760, 0.783] |  |
| Substance Abuse Problem |  |  |
| Yes | 1.136 [1.120, 1.152] | <0.0001 |
| No/ Not Reported (ref) | — |  |
| Service Site |  |  |
| Community-Based Program | — | <.0001 |
| Justice System Institution | 45.115 [42.543, 47.828] |  |
| Residential Treatment Center | 4.570 [4.347, 4.807] |  |
| State Psych Hospital | 3.988 [3.883, 4.096] |  |
| Other Psych inpatient | 1.724 [1.696, 1.752] |  |
| SMI/SED |  |  |
| SMI | 1.207 [1.185, 1.229] | <0.0001 |
| SED and/or at risk for SED | 1.713 [1.674, 1.752] |  |
| Not SMI/SED (ref) | — |  |
| NUMMHS |  |  |
| 0 | 1.495 [1.470, 1.520] | <0.0001 |
| 1 | 1.114 [1.105, 1.123] |  |
| 2 | — |  |
| 3 | 1.007 [0.992, 1.022] | 0.3302 |
| Behavioral/Cognitive Disorder |  |  |
| Yes | 0.83 (0.80–0.87) | 0.3443 |
| No (ref) | — |  |

The interaction analysis revealed several significant effect modifications in the relationship between behavioral and cognitive disorders and residential status. Participants aged 0–17 (AOR = 0.855, 95% CI: 0.802–0.912) and 45+ (AOR = 0.846, 95% CI: 0.828–0.865) had lower odds of discharge to non-private residence compared to those aged 18–44. Females had slightly higher odds than males (AOR = 1.087, 95% CI: 1.064–1.111). Hispanic (AOR = 1.181, 95% CI: 1.139–1.224), non-Hispanic Asian (AOR = 1.314, 95% CI: 1.146–1.507), non-Hispanic Black (AOR = 1.032, 95% CI: 1.007–1.058), and multiracial/other race individuals (AOR = 1.091, 95% CI: 1.037–1.147) were more likely to be discharged to non-private residences than non-Hispanic Whites.

Employment and education also showed significant interaction effects. Unemployed individuals had lower odds of non-private residence (AOR = 0.845, 95% CI: 0.814–0.877), and those without a high school diploma had reduced odds (AOR = 0.927, 95% CI: 0.906–0.949), while beyond high school education was not significant (AOR = 1.001, 95% CI: 0.975–1.028). Substance use was associated with increased odds of non-private residence (AOR = 1.056, 95% CI: 1.034–1.078). Compared to community-based programs, discharges from justice system institutions (AOR = 1.272, 95% CI: 1.121–1.444), residential treatment centers (AOR = 1.169, 95% CI: 1.086–1.258), and other psychiatric inpatient sites (AOR = 1.105, 95% CI: 1.060–1.151) had higher odds, while state psychiatric hospitals had lower odds (AOR = 0.803, 95% CI: 0.764–0.844). Participants with no prior mental health diagnosis (AOR = 1.495, 95% CI: 1.443–1.548), one diagnosis (AOR = 1.114, 95% CI: 1.088–1.141), and three diagnosis (AOR = 1.036, 95% CI: 1.002–1.071) all had higher odds of discharge to non-private residence.

**Table 3:** Interaction Effects model demonstrating the association between behavioral/cognitive disorders and Covariates on residential status outcomes at discharge (AOR, 95% CI)

| Variable | AOR (95% CI) | P-value |
| --- | --- | --- |
| Age ( in years) |  |  |
| 0-17 | 0.855 [0.802 – 0.912] | <0.0001 |
| 18-44 (ref) | — |  |
| 45+ | 0.846 [0.828 – 0.865] |  |
| Gender |  |  |
| Male (ref) | — | <0.0001 |
| Female | 1.087 [1.064 – 1.111] |  |
| Race/ethnicity |  |  |
| Hispanic | 1.181 [1.139 – 1.224] | <0.0001 |
| Non-Hispanic Native American | 1.059 [0.970 – 1.156] | 0.2007 |
| Non-Hispanic Asian | 1.314 [1.146 – 1.507] | <0.0001 |
| Non-Hispanic Black | 1.032 [1.007 – 1.058] | 0.0134 |
| Non-Hispanic Pacific Island | 1.201 [0.937 – 1.540] | 0.1582 |
| Non-Hispanic White (ref) | — | — |
| Other race/ Two or more | 1.091 [1.037 – 1.147] | 0.0002 |
| Employment Status |  |  |
| Employed (ref) | — | <0.0001 |
| Unemployed | 0.845 [0.814 – 0.877] |  |
| Education |  |  |
| No Diploma/Under 12th | 0.927 [0.906 – 0.949] | <0.0001 |
| Highschool Diploma/GED | — |  |
| Beyond Highschool | 1.001 [0.975 – 1.028] | 0.9194 |
| Substance Abuse Problem |  |  |
| Yes | 1.056 [1.034 – 1.078] | <0.0001 |
| No/ Not Reported (ref) | — |  |
| Service Site |  |  |
| Community-Based Program | — | <.0001 |
| Justice System Institution | 1.272 [1.121 – 1.444] |  |
| Residential Treatment Center | 1.169 [1.086 – 1.258] |  |
| State Psych Hospital | 0.803 [0.764 – 0.844] |  |
| Other Psych inpatient | 1.105 [1.060 – 1.151] |  |
| SMI/SED |  |  |
| SMI | 1.112 [1.078 – 1.146] | <0.0001 |
| SED and/or at risk for SED | 0.944 [0.887 – 1.004] | 0.0964 |
| Not SMI/SED (ref) | — |  |
| NUMMHS |  |  |
| 0 | 1.495 [1.443 – 1.548] | <0.0001 |
| 1 | 1.114 [1.088 – 1.141] |  |
| 2 | — | — |
| 3 | 1.036 [1.002 – 1.071] | 0.0192 |

## DISCUSSION

This study investigated the relationship between residential status and behavioral or cognitive disorders in the United States, using the 2022 Mental Health Client-Level Data (MH-CLD) (Mental Health Client-Level Data, 2024). The results demonstrated that those with behavioral or cognitive disorders are significantly less likely to reside in private residences compared to experiencing homelessness or living in alternative settings, such as foster care or correctional facilities. Covariates such as education, gender, employment status, and socioeconomic factors were found to significantly influence residential outcomes, with higher education and employment levels improving the likelihood of residential stability. These findings align with prior literature emphasizing the vulnerabilities of those with mental health conditions in institutional and unstable living arrangements (Breedvelt et al., 2019; Riske-Morris et al., 2023), while providing new insights into the specific demographic and socioeconomic predictors that affect residential outcomes.

The interpretation of these findings highlights the broader impact of socioeconomic disparities. For instance, the strong relationship between higher education and private residence stability underscores the importance of access to resources, while the higher prevalence of homelessness among children with disorders emphasizes gaps in social safety nets (World Health Organization, 2022). Gender differences in outcomes, where females are less likely to experience homelessness than males, might reflect differential access to social support services or cultural dynamics (Riske-Morris et al., 2023). Additionally, the partial mediation of these outcomes by socioeconomic factors indicates complex mechanisms involving support, mental health literacy, and systemic inequalities (Breedvelt et al., 2019).

Substance use emerged as a significant factor influencing residential stability, with those experiencing substance use problems being 20.2% less likely to reside in private residences compared to those without such issues. This shows the widespread impact of substance use on housing outcomes across populations, reflecting both individual-level challenges and broader systemic barriers. Substance use often co-occurs with other mental health conditions, creating a cycle of instability that limits access to secure and supportive housing. Addressing these issues through comprehensive interventions strategies, including integrated mental health, substance use treatment, and housing support programs, is important when trying to improve residential outcomes and overall well-being for affected individuals.

While this study contributes valuable insights, several limitations should be noted. The cross-sectional design limits causal inferences, as associations are captured at a single point in time (Kwan & Rickwood, 2015). The reliance on categorized and self-reported data may introduce inaccuracies in diagnostic and residential classifications (Mental Health Client-Level Data, 2024). Furthermore, the exclusion of secondary and tertiary diagnoses may underestimate the broader mental health burden and any missing data may slightly affect the robustness of findings. Finally, the absence of longitudinal follow-up limits the understanding of transitions in residential stability over time, a point noted as a gap in prior literature as well (Breedvelt et al., 2019).

These findings suggest important implications for public health and policy. Interventions should focus on addressing socioeconomic barriers, such as caregiver education and employment, to improve residential stability for children with mental health disorders (Riske-Morris et al., 2023; World Health Organization, 2022). Housing programs integrated with mental health services could provide holistic support, addressing both immediate and underlying needs. Future research should employ longitudinal designs to capture the dynamic relationship between mental health and residential outcomes, while also addressing unmeasured variables such as cultural influences and systemic barriers to care (Kwan & Rickwood, 2015; Riske-Morris et al., 2023).

## CONCLUSION

This study highlights the significant associations between behavioral and cognitive disorders and residential status amongst different age groups, emphasizing the role of socioeconomic and demographic factors in shaping housing outcomes. The findings underscore the urgent need for targeted interventions that address housing stability and its interplay with mental health, particularly among vulnerable populations, like those dealing with substance use problems. By shedding light on these critical associations, this research provides a foundation for future studies and practical interventions aimed at reducing disparities and improving residential outcomes for all with mental health conditions.

## Data Availability

Substance Abuse and Mental Health Services Administration (https://www.samhsa.gov/data/report/2022-mental-health-client-level-data-annual-report)

